# Spatial and temporal invasion dynamics of the 2014-2017 Zika and chikungunya epidemics in Colombia

**DOI:** 10.1101/2020.09.11.20189811

**Authors:** Kelly Charniga, Zulma M. Cucunubá, Marcela Mercado, Franklyn Prieto, Martha Ospina, Pierre Nouvellet, Christl A. Donnelly

## Abstract

Zika virus (ZIKV) and chikungunya virus (CHIKV) were recently introduced into the Americas resulting in significant disease burdens. Understanding their spatial and temporal dynamics at the subnational level is key to informing surveillance and preparedness for future epidemics. We analyzed anonymized line list data on approximately 105,000 Zika virus disease and 412,000 chikungunya fever suspected and laboratory-confirmed cases during the 2014-2017 epidemics. We first determined the week of invasion in each city. Out of 1,122, 288 cities met criteria for epidemic invasion by ZIKA and 338 cities by CHIKV. We estimated that the geographic origin of both epidemics was located in Barranquilla, north Colombia. Using gravity models, we assessed the spatial and temporal invasion dynamics of both viruses to analyze transmission between cities. Invasion risk was best captured when accounting for geographic distance and intermediate levels of density dependence. Although a few long-distance invasion events occurred at the beginning of the epidemics, an estimated distance power of 1.7 (95% CrI: 1.5-2.0) suggests that spatial spread was primarily driven by short-distance transmission. Cities with large populations were more likely to spread disease than cities with smaller populations. Similarities between the epidemics included having the same estimated geographic origin and having the same five parameters estimated in the best-fitting models. ZIKV spread considerably faster than CHIKV.

**Author summary:** Understanding the spread of infectious diseases across space and time is critical for preparedness, designing interventions, and elucidating mechanisms underlying transmission. We analyzed human case data from over 500,000 reported cases to investigate the spread of the recent Zika virus (ZIKV) and chikungunya virus (CHIKV) epidemics in Colombia. Both viruses were introduced into northern Colombia. We found that intermediate levels of density dependence best described transmission and that transmission mainly occurred over short distances. Our results highlight similarities and key differences between the ZIKV and CHIKV epidemics in Colombia, which can be used to anticipate future epidemic waves and prioritize cities for active surveillance and targeted interventions.

## Introduction

The global burden of disease due to arboviral infections is substantial and continues to increase [1]. Chikungunya virus (CHIKV) is an alphavirus that is transmitted to people primarily by *Aedes* mosquitoes [2]. Cases of chikungunya fever, the disease caused by CHIKV, were first reported in the Americas in December 2013 [3]. Within a year, over one million cases were reported in the region, including severe cases and deaths [4]. Zika virus (ZIKV) is a flavivirus that is also spread by *Aedes* mosquitoes. Cases of ZIKV disease were first reported in Brazil in May 2015. From Brazil, ZIKV spread widely throughout Latin America and the Caribbean. In February 2016, the World Health Organization declared ZIKV a Public Health Emergency of International Concern [5]. While symptoms of the two diseases typically include fever, rash, and joint pain, ZIKV infection during pregnancy has been associated with severe birth defects in fetuses and newborns [6], and chikungunya fever can cause chronic joint pain that lasts months or years [3]. There are currently no approved drugs to treat or prevent ZIKV disease or chikungunya fever, although several vaccine candidates are under investigation [7, 8].

Previously, the spatial and temporal spread of ZIKV [9-11] and CHIKV [12-14] in the Americas has been studied separately. However, the viruses share common vectors and were both introduced into apparently immunologically naïve populations. Studying these diseases together in the same country may help elucidate similarities and differences between the two. Here, a suite of gravity models was fitted to analyze transmission between cities in Colombia. The invasion dynamics of both viruses were examined as well as the extent to which inter-city transmission depended on distance, population sizes of invaded and susceptible cities, and the infectivity of ZIKV and CHIKV.

## Results

### Temporal and spatial patterns in invasion weeks

Out of 1,122 cities in Colombia, week of invasion was determined for 338 cities for CHIKV and 288 cities for ZIKV. Invasion weeks ranged from the week ending May 31, 2014 to that ending September 19, 2015 for CHIKV and from the week ending August 8, 2015 to that ending March 26, 2016 for ZIKV. The time for the diseases to invade 50% of cities ever affected was shorter for ZIKV compared to CHIKV (Table 1), and while onset times for ZIKV tended to cluster within five months (from September to January), 90% of onset times for CHIKV clustered within nine months (between September and May). For cities that experienced epidemics of both CHIKV and ZIKV (n = 205), invasion weeks were significantly positively correlated (Pearson’s correlation coefficient 0.45, p < 0.0001). Both epidemics were first recorded in northern Colombia and spread from there. Early foci of disease were also present in the central parts of the country (Fig 1).

**Table 1.**
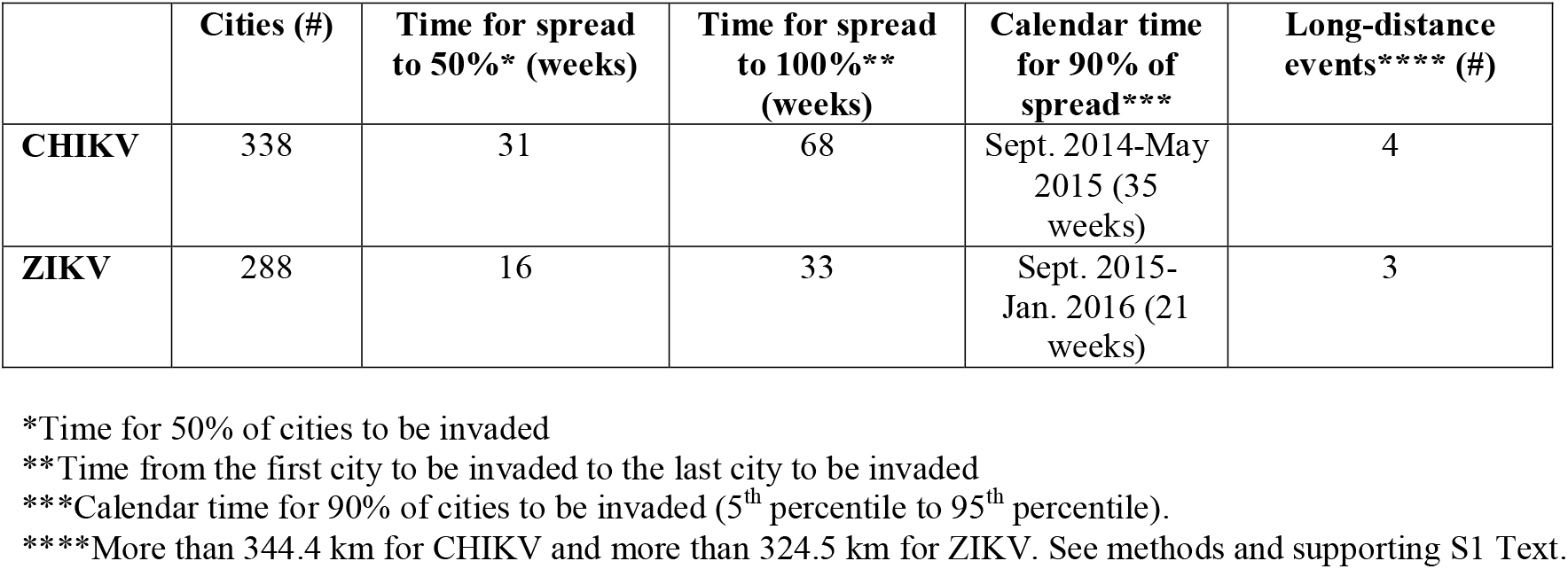
Epidemiological characteristics of CHIKV and ZIKV epidemics in Colombia.

|  | Cities (#) | Time for spread to 50%* (weeks) | Time for spread to 100%** (weeks) | Calendar time for 90% of spread*** | Long-distance events**** (#) |
| --- | --- | --- | --- | --- | --- |
| **CHIKV** | 338 | 31 | 68 | Sept. 2014-May 2015 (35 weeks) | 4 |
| **ZIKV** | 288 | 16 | 33 | Sept. 2015-Jan. 2016 (21 weeks) | 3 |

\*Time for 50% of cities to be invaded
\*\*Time from the first city to be invaded to the last city to be invaded
\*\*\*Calendar time for 90% of cities to be invaded (5<sup>th</sup> percentile to 95<sup>th</sup> percentile).
\*\*\*\*More than 344.4 km for CHIKV and more than 324.5 km for ZIKV. See methods and supporting S1 Text.

**Fig 1.**
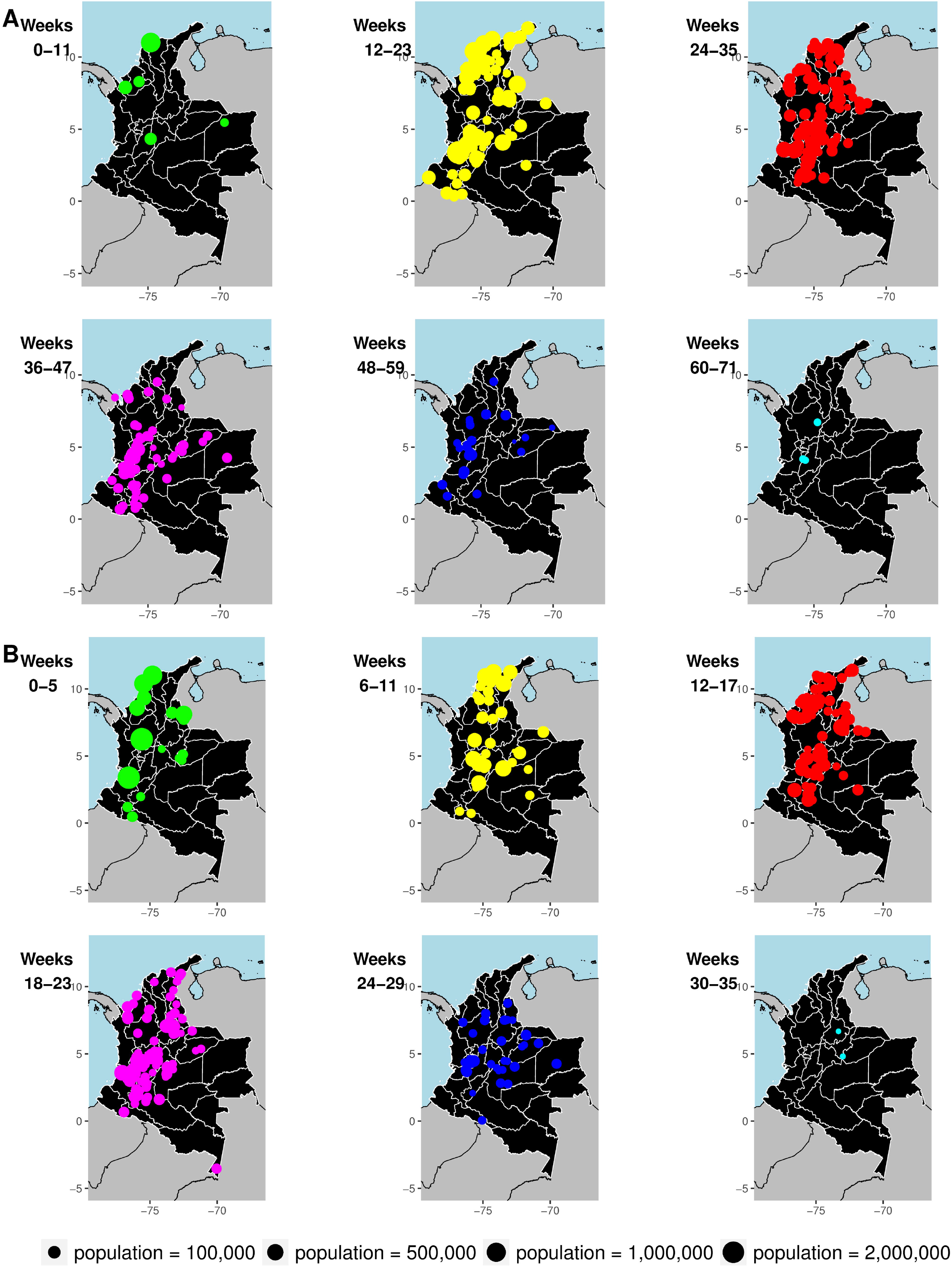
Geographic patterns of invasion times in studied cities in Colombia based on first reported cases. Invasion times are shown by 12-week period for (A) CHIKV and 6-week period for (B) ZIKV. Each circle represents a city, and the size of the circle is proportional to population size. Each panel shows only cities newly invaded during the time period indicated in the upper left-hand-corner. The island of San Andrés is not shown but was invaded by CHIKV in week 21 and by ZIKV in week 0.

### Geographic origin of epidemics

For both epidemics, the estimated origin was Barranquilla, Colombia’s fourth most populated city located on the Caribbean coast (Fig 2). Fig 3 shows the spread of reported cases during the epidemics, and S1-S2 Movies show the monthly incidence per 100,000 population by first administrative unit (department).

**Fig 2.**
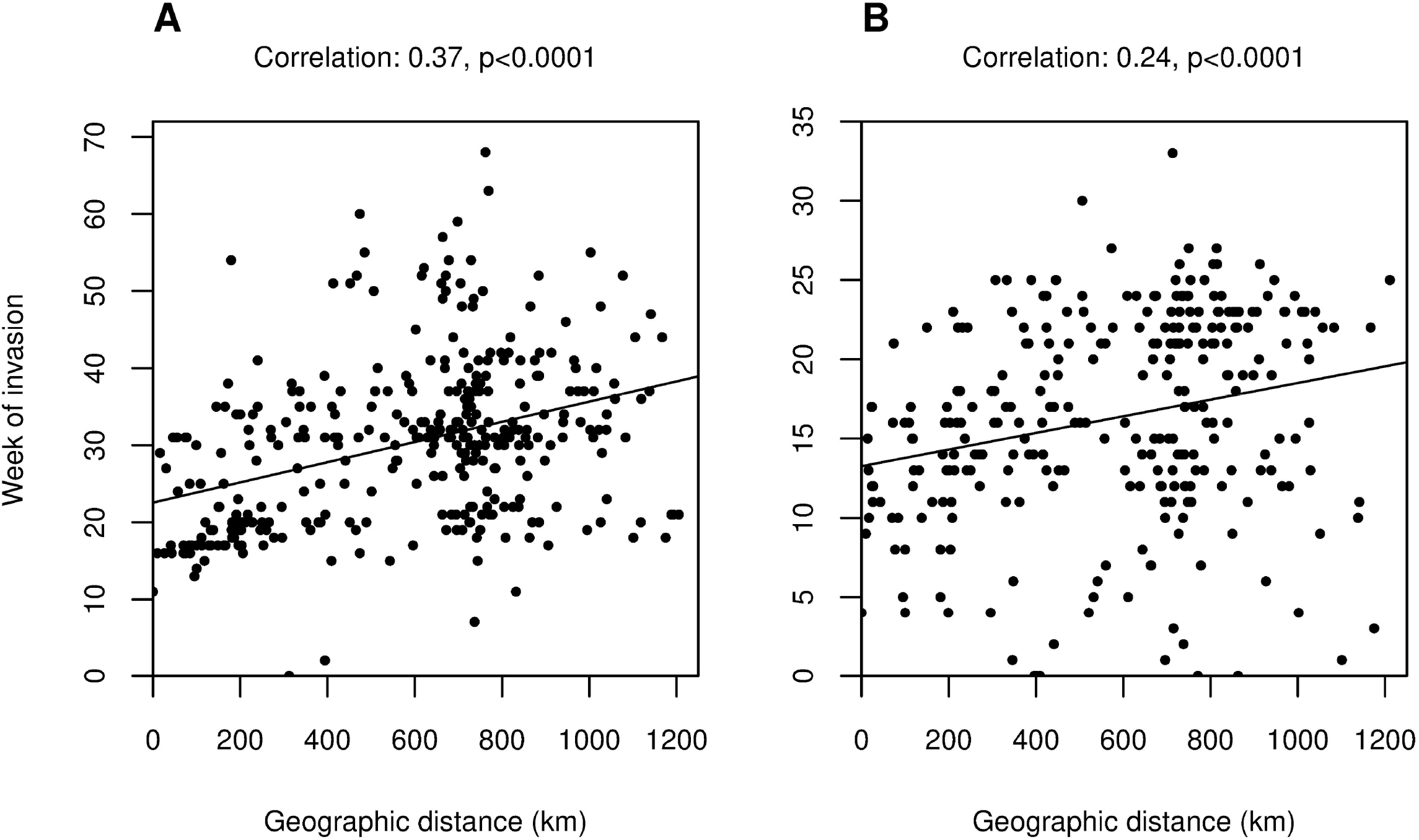
Correlations between city invasion times and geographic distance from first invaded cities. Week of invasion is shown on the y-axis for both plots. These weeks are correlated as all cities ever invaded against (A) geographic distance from the most likely origin of CHIKV in Colombia, Barranquilla and (B) geographic distance from the most likely origin of ZIKV in Colombia, also Barranquilla. Pearson’s correlation coefficients and significance are shown above each plot.

**Fig 3.**
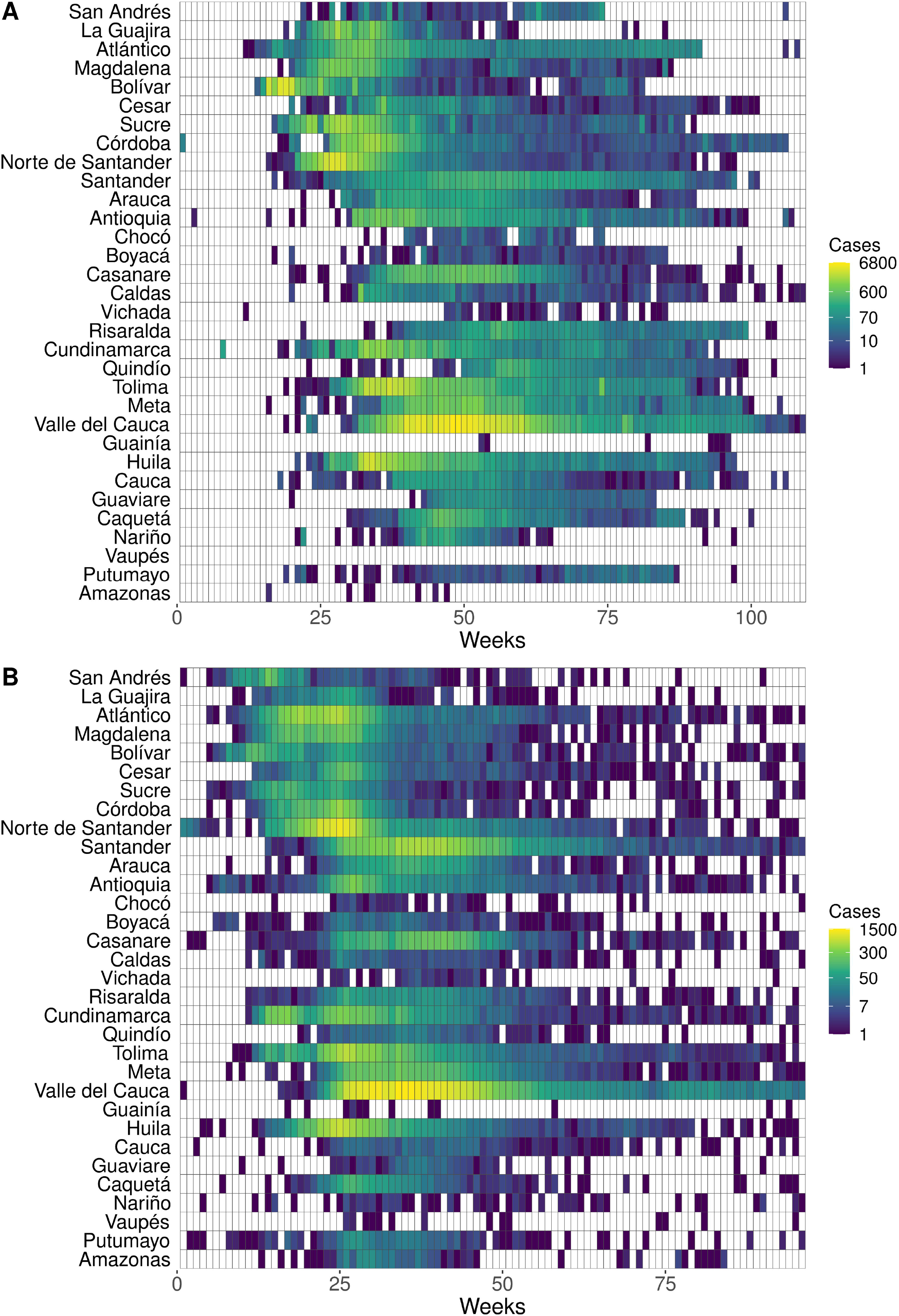
Heatmaps showing the spatial and temporal spread of CHIKV and ZIKV in Colombia. Population-weighted centroids were used to rank departments in order from North to South. Colors across rows represent the number of cases of (A) chikungunya fever and (B) ZIKV disease for each department. Weeks are plotted on the x-axis starting from the first week cases were reported to the last week cases were reported. Dates for (A) range from the week ending June 7, 2014 to that ending July 9, 2016, and dates for (B) range from the week ending August 15, 2015 to that ending June 17, 2017. White rectangles are weeks with zero reported cases.

### Long-distance transmission events

Four long-distance transmission events were identified for CHIKV and three were identified for ZIKV. For CHIKV, the affected cities were Girardot, Cundinamarca; La Primavera, Vichada; and Mocoa and Puerto Asís in the department of Putumayo. For ZIKV, the affected cities were Barranquilla, Atlántico; Tauramena, Casanare; and Mocoa, Putumayo. Two of these six cities are department capitals. All of these events occurred early in the epidemics, within the first 15% of cities invaded (methods and supporting S1 Text). It is important to note that the methods for estimating the epidemic origin and long-distance transmission events are independent of one another. In the case of ZIKV, for example, Barranquilla is estimated as both the epidemic origin and a long-distance transmission event.

### Models of spread

Although geographic distance fit the epidemic data better than travel time between cities for ZIKV, the fits of the CHIKV model variants were indistinguishable (supporting S1 Text). Tables 2 and 3 show the model results for the geographic distance variant.

**Table 2.** Parameter estimates for six models of CHIKV in Colombia for 338 cities.

| | Distance-only model | Density-dependent population model | Density-independent population model | Estimated density dependence population model | Full (infectivity) model | Infectivity model with $\mu$ set to 0* |
| --- | --- | --- | --- | --- | --- | --- |
| DIC | 2521.0 | 2448.4 | 2403.0 | 2402.6 | 2336.1 | **2335.1** |
| $\gamma$ (distance power) | 1.03<br>(0.83, 1.21) | 1.02<br>(0.84, 1.17) | 1.46<br>(1.22, 1.70) | 1.49<br>(1.25, 1.72) | 1.68<br>(1.43, 1.90) | **1.68**<br>**(1.44, 1.90)** |
| $\mu$ (susceptible population) | 0 | 0.076<br>(0.003, 0.286) | 0.036<br>(0.001, 0.163) | 0.030<br>(0.001, 0.141) | 0.061<br>(0.002, 0.207) | **0** |
| $v$ (invaded population) | 0 | 0.45<br>(0.34, 0.55) | 0.54<br>(0.42, 0.65) | 0.53<br>(0.41, 0.64) | 0.64<br>(0.52, 0.76) | **0.65**<br>**(0.53, 0.76)** |
| $\varepsilon$ (spatial interaction) | 0 | 0 | 1 | 0.86<br>(0.68, 1.05) | 0.84<br>(0.69, 1.00) | **0.83**<br>**(0.69, 0.98)** |
| $\phi$ (infectivity) | 0 | 0 | 0 | 0 | 0.34<br>(0.25, 0.46) | **0.35**<br>**(0.25, 0.48)** |
| $\beta$ (intensity) | 0.15<br>(0.05, 0.37) | 0.08<br>(0.03, 0.19) | 0.26<br>(0.20, 0.31) | 0.30<br>(0.22, 0.43) | 0.21<br>(0.11, 0.35) | **0.24**<br>**(0.13, 0.39)** |

Posterior medians and 95% credible interval presented for each parameter. Bold indicates the best-fitting model.
\*When $\mu$ is set to 0, this means that cities with large populations have the same risk of being invaded as cities with small populations.

**Table 3.**
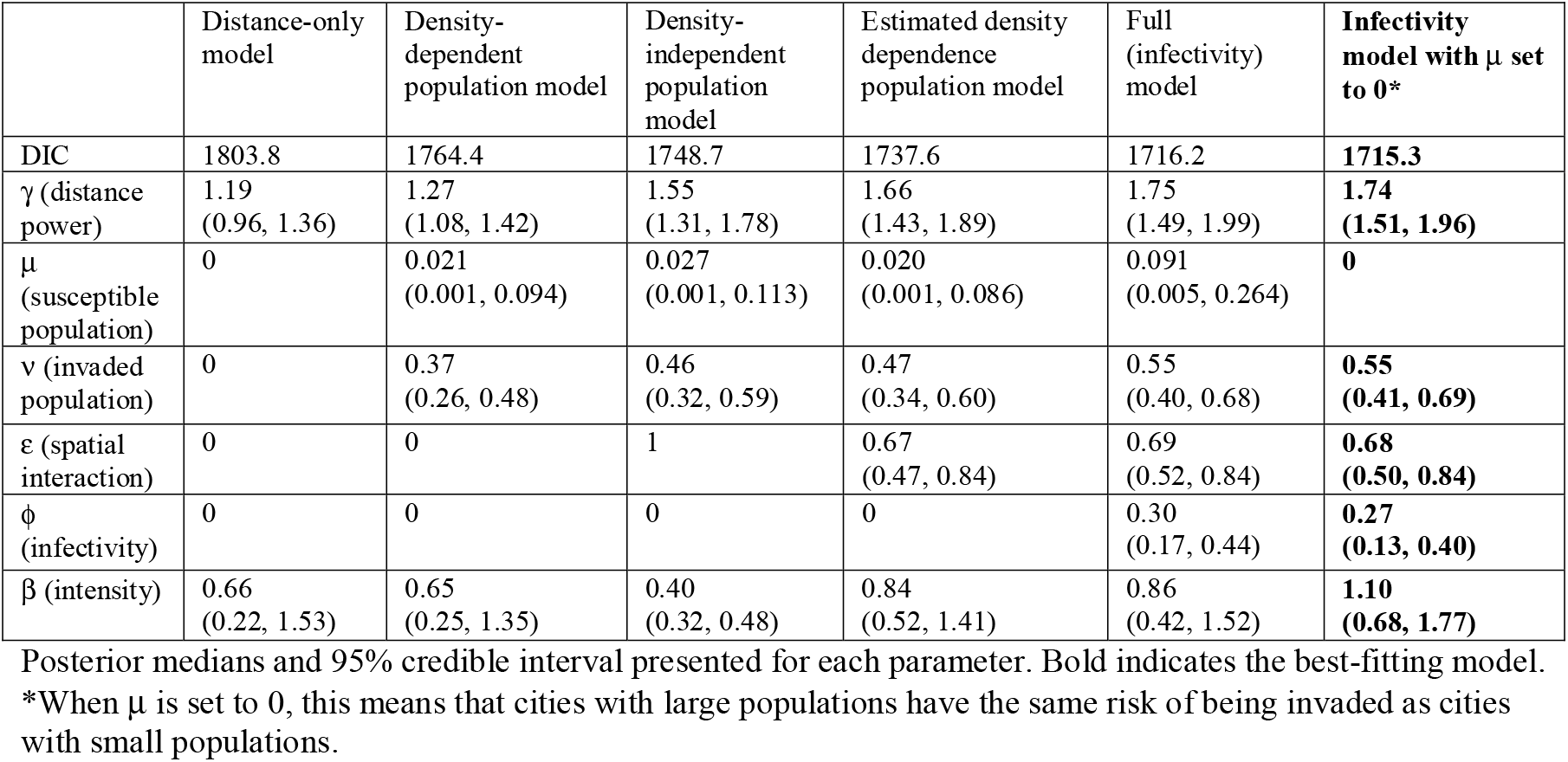
Parameter estimates for six models of ZIKV in Colombia for 288 cities.

| | Distance-only model | Density-dependent population model | Density-independent population model | Estimated density dependence population model | Full (infectivity) model | Infectivity model with $\mu$ set to 0* |
| --- | --- | --- | --- | --- | --- | --- |
| DIC | 1803.8 | 1764.4 | 1748.7 | 1737.6 | 1716.2 | **1715.3** |
| $\gamma$ (distance power) | 1.19<br>(0.96, 1.36) | 1.27<br>(1.08, 1.42) | 1.55<br>(1.31, 1.78) | 1.66<br>(1.43, 1.89) | 1.75<br>(1.49, 1.99) | **1.74**<br>**(1.51, 1.96)** |
| $\mu$ (susceptible population) | 0 | 0.021<br>(0.001, 0.094) | 0.027<br>(0.001, 0.113) | 0.020<br>(0.001, 0.086) | 0.091<br>(0.005, 0.264) | **0** |
| $v$ (invaded population) | 0 | 0.37<br>(0.26, 0.48) | 0.46<br>(0.32, 0.59) | 0.47<br>(0.34, 0.60) | 0.55<br>(0.40, 0.68) | **0.55**<br>**(0.41, 0.69)** |
| $\varepsilon$ (spatial interaction) | 0 | 0 | 1 | 0.67<br>(0.47, 0.84) | 0.69<br>(0.52, 0.84) | **0.68**<br>**(0.50, 0.84)** |
| $\phi$ (infectivity) | 0 | 0 | 0 | 0 | 0.30<br>(0.17, 0.44) | **0.27**<br>**(0.13, 0.40)** |
| $\beta$ (intensity) | 0.66<br>(0.22, 1.53) | 0.65<br>(0.25, 1.35) | 0.40<br>(0.32, 0.48) | 0.84<br>(0.52, 1.41) | 0.86<br>(0.42, 1.52) | **1.10**<br>**(0.68, 1.77)** |

Posterior medians and 95% credible interval presented for each parameter. Bold indicates the best-fitting model.
\*When $\mu$ is set to 0, this means that cities with large populations have the same risk of being invaded as cities with small populations.

For both epidemics, the best model (based on the lowest Deviance Information Criterion, DIC) included the following parameters: a distance power, power for invaded city population size, density dependence, infectivity, and transmission intensity. In both instances, the population size of the susceptible city appeared uncorrelated with the invasion dynamics. The estimated distance power, γ, was 1.68 (95% CrI: 1.44-1.90) for CHIKV and 1.74 (95% CrI: 1.51-1.96) for ZIKV. The credible intervals for γ overlap, so we cannot exclude the possibility that the relationship with distance is the same for both viruses. Both models also estimated intermediate levels of density dependence (ε between 0 and 1).

For each city, we evaluated the predicted invasion week given the observed invasion weeks in other cities up to that time. The best-fitting models predicted the distribution of the local start of epidemics well (Fig 4). Excluding cities that were invaded in week 0, for CHIKV 304 out of 337 cities (90%, 95% CI: 87-93%) lie within the 95% interval of their expected distribution, and for ZIKV 268 out of 283 cities (95%, 95% CI: 91-97%) lie within the 95% interval of their expected distribution. Cities that fell outside of these intervals tended to be invaded at the beginning or the end of the epidemics (supporting S1 Text). The best-fitting ZIKV model captured the shape of the observed invasion week distribution well. In contrast, the best-fitting CHIKV model did not capture the shape well at the end of the epidemic. Cities invaded late in the CHIKV epidemic (week 53 or later) had smaller population sizes and fewer cases compared to cities invaded earlier (up to week 52) (Wilcoxon rank sum tests for population size: W = 885, p = 0.004 and for cumulative case numbers: W = 900, p = 0.005).

**Fig 4.**
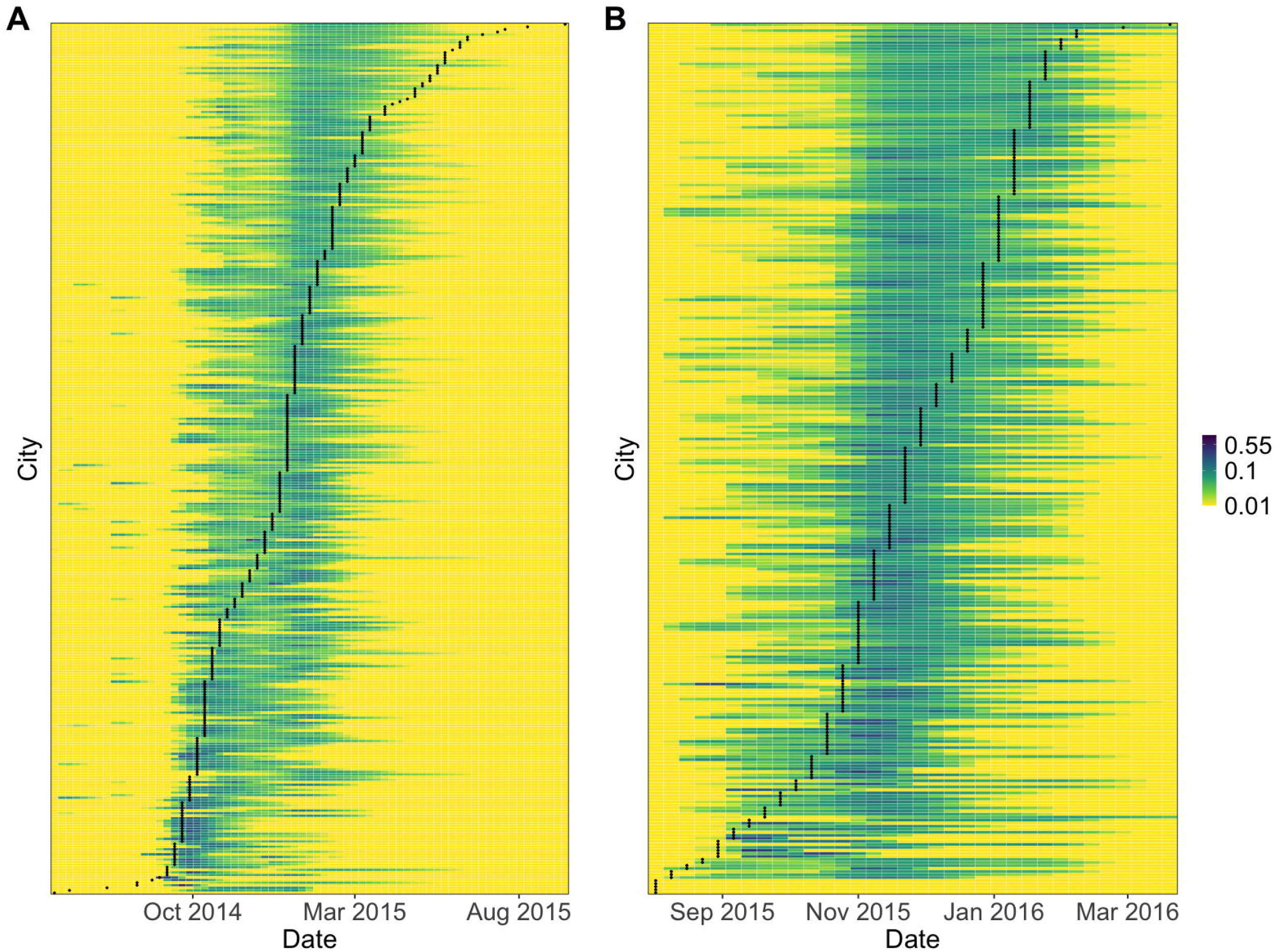
Probability distribution of invasion weeks.

The panels show the estimated probability distributions of invasion week for each city (colored lines) for (A) CHIKV and (B) ZIKV based on the observed start of invasion in other cities up to that time. The calculations were performed using the median parameter estimates from the posterior distributions of the best-fitting models for CHIKV and ZIKV. The black lines show the observed invasion week based on the first reported cases in each city. Values plotted as 0.01 represent probabilities of 0.01 or less.

Simulated epidemics from the best-fitting model for each virus were consistent with the observed epidemic in terms of the number of invaded cities over time (Fig 5). For CHIKV, we observed half of the cities invaded by week 31 of the epidemic, while 1,000 simulations predicted half of the cities invaded by week 34.4 on average (min. 17, mode 27, max. 66). For ZIKV, we observed half of the cities invaded by week 16, while simulations predicted 15.7 on average (min. 11, mode 15, max. 23).

**Fig 5.**
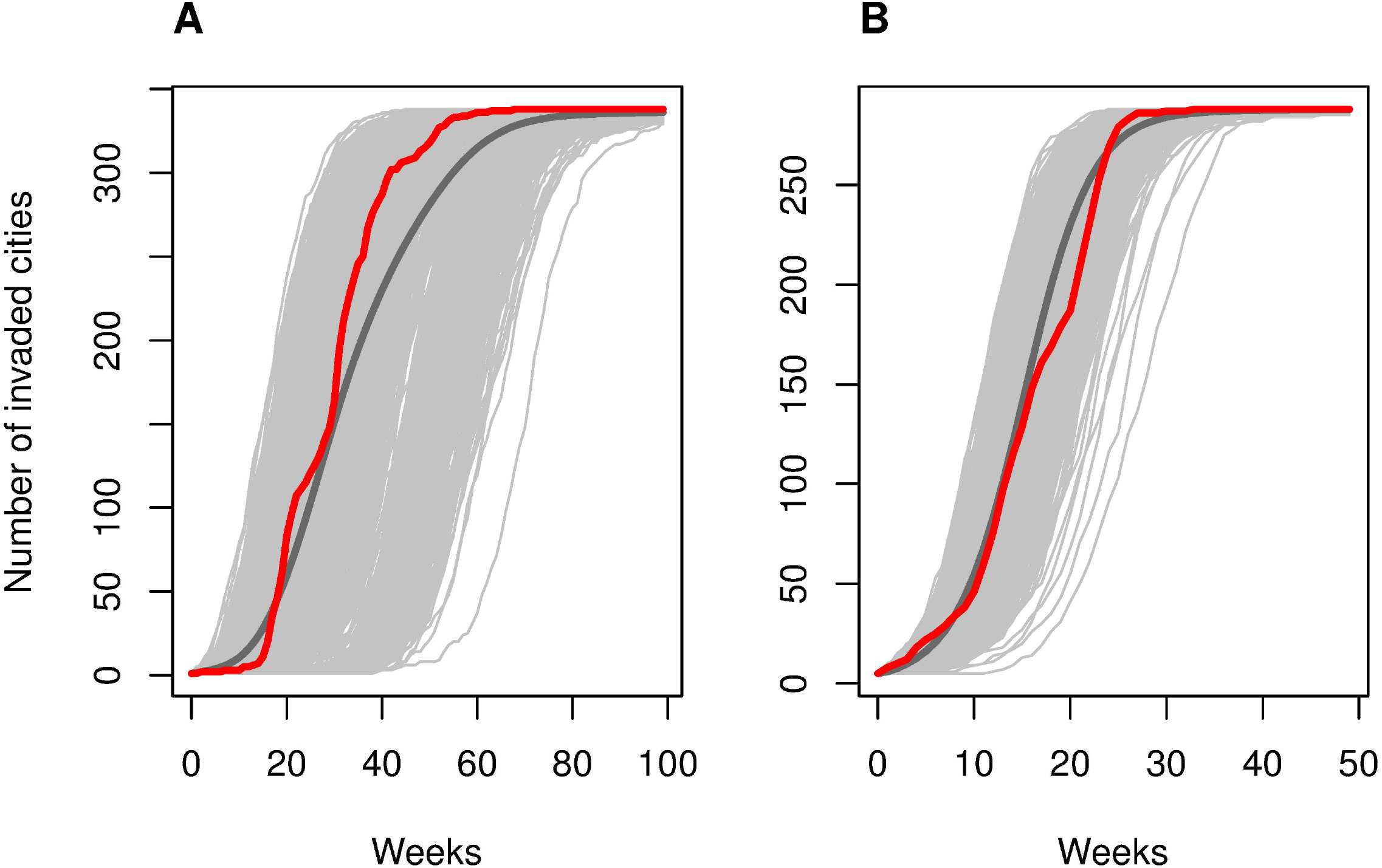
Epidemic invasion simulations. Simulated invasion time (as week of first reported cases) for (A) CHIKV and (B) ZIKV from the best-fitting models. Simulated epidemics are shown in light gray. The dark gray lines are the average across the 1,000 simulations. The red lines show the observed number of cities that first reported cases in each week.

Epidemic simulations showed that model fits for ZIKV slightly improved visually when the cut-off for invasion was set at 30 reported cases rather than 20 (results not shown).

Results of models fitted to all 1,122 cities in Colombia are presented in the supporting S1 Text. When all cities were included, as expected the transmission intensity estimate was much lower, and the epidemic simulations showed a very delayed and prolonged epidemic compared to the observed incidence of invaded cities.

## Discussion

Similarities and key differences in the space-time dynamics of the CHIKV and ZIKV epidemics in Colombia were identified using gravity models. Spatial invasion of both epidemics likely began in the north, where the Andes Mountains serve as a natural barrier to human movement. Although the best-fitting models for both viruses included the same five parameters, the ZIKV epidemic spread twice as fast as the CHIKV epidemic. Our parameter estimates for γ, ν, ε, and ϕ are consistent with those obtained in studies of seasonal and pandemic influenza spread [15, 16].

Geographic distance was the preferred distance metric to describe spread of ZIKV rather than estimated travel time between cities. For CHIKV, both distance metrics performed similarly. Although Viboud et al. found that work commutes better described the spread of seasonal influenza in the U.S. compared to geographic distance, Charu et al. found that geographic distance outperformed models with work commutes and models with air traffic [15, 17]. Geographic distance was also a better predictor of CHIKV spread in the Caribbean region than air traffic [12].

The estimated power of the effect of distance on spread, γ, with geographic distance was about 1.7 (Tables 1-2), indicating that transmission was dominated by short-distance interactions. Slightly higher estimates of about 2.0 were obtained for models using travel time between cities rather than geographic distance in 337 and 287 cities for which travel time data were available (supporting S1 Text). We expected similar estimates of γ for CHIKV and ZIKV because they were spread by the same vectors in the same geographic area. Using geolocated genotype and serotype data, Salje et al. found evidence that in Bangkok, Thailand, transmission of dengue virus, another arbovirus spread by *Aedes* mosquitoes, is highly focal, with the majority of infection events occurring near the home [18].

A range of estimates of γ have been reported in the literature. Gog et al. reported 2.6 (95% CI: 2.3-2.8) for 2009 pandemic influenza in the U.S., and Charu et al. reported a median of 2.2 (range 2.1-2.7 with standard deviations between 0.13 and 0.33) across seven influenza seasons in the U.S. [15, 19]. However, Eggo et al. reported lower values of 1.2 and 0.86 for 1918 pandemic influenza in England and Wales and in the U.S., respectively [16]. Differences could be attributed, at least in part, to data being aggregated at different spatial scales; fewer data points in an area will lead to lower estimates of the distance power because locations are farther apart.

For the best-fitting CHIKV and ZIKV models, we obtained similar estimates for the estimated power for the effect of invaded city population size v, indicating that cities with large populations are more likely to spread disease than cities with smaller populations. Gog et al. did not include this parameter, and in Eggo et al., it was not selected in their best-fitting England and Wales model.

For both CHIKV and ZIKV, we accepted the null hypotheses that the estimated powers (μ) for the effects of susceptible city population size were 0. This means that cities with large populations have the same risk of being invaded as cities with small populations. In contrast, low, but significant, estimates of μ were reported by Gog et al. (0.27, 95% CI: 0.11-0.44) and Eggo et al. (0.40, 95% CrI: 0.25-0.54) for seasonal and pandemic influenza, respectively.

Intermediate levels of density dependence best described transmission (ε for CHIKV 0.83, 95% CrI: 0.69-0.98 and ZIKV 0.68, 95% CrI: 0.50-0.84). In other words, connectivity somewhat depended on the number and size of neighboring populations. For influenza in the U.S., Gog et al. and Charu et al. reported estimates of ε close to 1 [15, 19]. Eggo et al. reported ε close to 1 for influenza in England and Wales but also found that a density-dependent model (ε = 0) fit the data best for influenza in the U.S. [16]. Similarly, Salje et al. found that dengue virus transmission in Bangkok, Thailand was consistent with density-dependent transmission (ε = 0) [18]. Differences in estimates could be due to differences in both coverage of datasets and spatial scales considered.

We obtained low estimates for the infectivity parameter ϕ for both CHIKV (0.35, 95% CrI: 0.25-0.48) and ZIKV (0.27, 95% CrI: 0.13-0.40) models. This suggests that cities with more reported cases were more infectious than cities with fewer reported cases. One reason for low, though significant, estimates could be because reported case incidence poorly reflects the true incidence of infection in a city. For example, if reporting rates vary over time or by location, cases reported to the surveillance system may not be a good proxy for actual infection incidence. Using mortality rate as a proxy for infectiousness, Eggo et al. reported a similar estimate of 0.24 (95% CrI: 0.03-0.47) for pandemic influenza in England and Wales [16].

Transmission intensity, represented by β, is the only parameter for which the credible intervals do not overlap between the two viruses. The estimated β for ZIKV is significantly higher than that for CHIKV, reflecting the faster spread of the ZIKV epidemic. Differences in transmission intensity could be related to the 2015-2016 El Niño weather phenomenon. Caminade et al. found that increased temperatures associated with El Niño created conditions across South America that were favorable for ZIKV transmission in 2015 [20].

The results presented here depend on estimates of invasion times in each city. We defined invasion time as the week before cases were first reported in each city. At the beginning of an outbreak, one or even a small number of reported cases in a city may not be sufficient to sustain chains of transmission resulting in spread to other cities; however, the first reported cases could be the indication of previously undetected transmission. A genomic epidemiological study found evidence that ZIKV had been circulating undetected in Colombia for five to eight months before the first cases were confirmed in September 2015 [21]. Moreover, because we modeled the city of likely infection rather than city of notification or residence (supporting S1 Text), it is possible that cities with better surveillance or healthcare infrastructure could have been the first to report cases in travelers returning from cities with no previous evidence of transmission.

Our results are robust to uncertainty in invasion weeks. We fitted the models using an alternative definition for invasion week (supporting S1 Text). Although model fits for CHIKV were slightly worse, ZIKV model fits were comparable. Reassuringly, parameter estimates were similar (supporting S1 Text).

Only cities that met thresholds for cumulative reported cases were included in the analysis. All other locations were treated as missing. Similar approaches have been employed in the study of seasonal and pandemic influenza [15, 16, 19]. Here, some unaffected cities were not invaded because they were not at risk (due to environmental factors). Of the cities that were at risk, some were invaded but others appeared to have escaped invasion by chance or other unexamined factors. Among cities that escaped invasion, there is also some probability that they had in fact been invaded but never reported cases. Populations of at least a sample of these cities warrant further investigation.

Ninety-nine percent of chikungunya fever cases and 95% of ZIKV disease cases were clinically confirmed, rather than laboratory confirmed. This could have led to misclassification, as dengue virus, CHIKV, and ZIKV were circulating simultaneously. Also, asymptomatic infection, mild illnesses, and limited access to healthcare likely resulted in underreporting. Problems with reporting and misdiagnoses may have affected the fit of the probability distribution of invasion week for cities invaded near the end of the CHIKV epidemic (Fig 4). Some of these late-invaded cities might have been invaded earlier but not reported cases to the surveillance system in a timely manner. Another possible explanation is that cases reported at the end of the CHIKV epidemic were actually misdiagnosed ZIKV disease cases.

A further limitation is that the model only incorporates one distance metric at a time. In reality, the spread of ZIKV and CHIKV was likely driven by a combination of air travel, land-based travel, and vector movement. The model also does not consider changes over time in reporting, human behavior, or transmission. These aspects could have changed during the epidemics, especially when the Public Health Emergency of International Concern was declared by the World Health Organization in February 2016 [5].

Another assumption of this model is that CHIKV and ZIKV were each introduced into Colombia only once. The results of two recent genomic studies suggest that this assumption is valid. Black et al. found evidence of two separate introductions of ZIKV into Colombia; however, the majority of cases were associated with a single introduction [21]. Similarly, Villero-Wolf et al. found evidence of only three introductions of CHIKV in Colombia, suggesting that most cases resulted from transmission within the country, rather than repeated travel-related importations [22].

The gravity model formulation used in this study works well retrospectively; however, more work is needed to understand why some cities appear to escape invasion. Until this issue is resolved, these methods have limited use for real-time forecasting of epidemics.

Future directions for this work include the use of this approach to understand the invasion dynamics of other epidemics. Further research should also focus on quantifying the relative contribution of human versus vector movement on spatial transmission. This would have broad implications for surveillance and control for other mosquito-borne epidemics such as dengue, Mayaro, and yellow fever.

## Materials and methods

### Data

We analyzed anonymized line list data on 105,152 ZIKV disease and 411,789 chikungunya fever suspected and laboratory-confirmed cases reported to Sivigila, Colombia’s national public health surveillance system, between 2014 and 2017. See supporting S1 Text for a full description of epidemiological and demographic data and data on human mobility proxies and elevation.

### Definition of invasion week

Out of 1,122 cities in Colombia, only cities with at least 20 cases of chikungunya fever were considered to have been “invaded” by CHIKV. Similarly, only cities with at least 30 cases of ZIKV disease were considered to have been “invaded” by ZIKV. Cities with case counts below these thresholds were not considered in the primary analysis. A higher threshold was used for ZIKV than for CHIKV based on our epidemic simulations. Invasion was defined as the week before cases were first reported in each city. We assumed a latent period of one week after which the city is considered infectious and can spread the infection to other cities. The supporting S1 Text includes details on and model fits to an alternative definition for invasion week.

### Potential sources of the epidemics

The cities in Colombia where the CHIKV and ZIKV epidemics most likely originated were identified. The method is based on the idea that epidemics spread radially from the origin, meaning that the relationship between invasion week and distance from the source is linear [15]. Assuming a single introduction of each virus into Colombia, the first 10% of invaded cities were considered as potential origins for the epidemics. Pearson’s correlation coefficient for the relationship between the city’s invasion week and its geographic distance to the origin was calculated for each potential origin. The most likely source was identified as the city with the highest such correlation coefficient (Fig 2).

### Long-distance transmission events

The number and location of long-distance transmission events of CHIKV and ZIKV were identified using the invasion week in each city. The method detects outliers in the distribution of pairwise distances between newly invaded cities and the set of infectious cities at the previous time step [15]. *C* is defined as the set of cities in the network. At time *t_j_*, *C* is divided into the set of infectious cities *I_tj_* and the set of susceptible cities *S_tj_*:

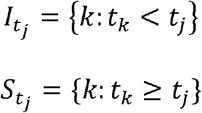

For city *j*, the minimum distance between city *j* and infectious cities in *I_tj_* was calculated as *d_j_*, the most likely route of invasion when the spatial dynamics are dominated by distance.

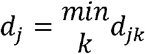

for *k* ∊ *I_tj_* We also calculated *D_j_*, the minimum distance between city *j* and any other city in the network:

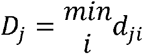

for *i* ∊ *C*. If the process were entirely spatial, cities would usually be invaded by nearby cities. Thus, the distance to the nearest city is approximated by *d_j_* − *D_j_* ≈ 0. For each city, *d_j_* − *D_j_* was calculated and those included in the 99^th^ percentile of the distribution of *d_j_* − *D_j_* were considered long-distance transmission events (supporting S1 Text).

### Gravity models of disease spread

We fitted gravity models to analyze transmission of CHIKV and ZIKV between cities that reported the minimum number of cases. These models have been used to study transmission of infectious diseases, such as measles [23], influenza [15-17, 19], vector-borne diseases [24], and cholera [25, 26], among others.

For each ZIKV and CHIKV separately, *N* cities have an invasion week, *t_i_*, which was defined as one week before cases were first reported in each city. Cities also have population size, *P_i_*, which is assumed to be constant over time and weekly case counts weighted by the generation time distribution, *c_i,t_* (supporting S1 Text). The generation time is the average time between the time of infection in a primary case and the time of infection in a secondary case infected by the primary case [27]. The geographic distance in km (or travel time in minutes) between invaded city *i* and susceptible city *j* is *d_ij_*. For geographic distance, we used the geodesic distance on an ellipsoid, which is the shortest path between two points accounting for the curvature of the Earth. Further details are given in the supporting S1 Text.

At each time point, a city can be either “susceptible,” “latently invaded,” or “infectious.” Once cities are invaded, they are latently invaded for one week and then infectious. After external seeding into Colombia occurs, we assumed no additional cases are imported from abroad. If a city is invaded in week *t_i_*, only Colombian cities that were infectious in the previous week could have spread the disease to that city. We assumed transmission parameters remain constant over time.

As in Eggo et al.[16], the force of infection, λ, represents the hazard of infection from an invaded city to a susceptible city. At time *t*, the force of infection from city *i* to city *j* can be defined as:

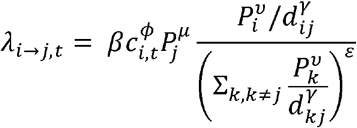

Exponents ν and μ are for population sizes of city *i* and *j*, respectively. The distance between cities is *d_ij_* and γ is the power parameter. β describes transmission intensity. ϕ captures the relationship between infectivity of a city and its weekly case count weighted by the generation time distribution. A value of ϕ = 1 indicates that the infectiousness of a city at time *t_i_* is proportional to the number of cases reported in that city weighted by the generation time distribution at time *t_i_* When ϕ = 0, infectiousness does not depend on the number of reported cases in the source city. Values of ϕ between 0 and 1 lead to infectiousness profiles that vary according to weekly case counts. Parameter ε characterizes the density dependence of the connection between a susceptible city and all invaded cities. When ε = 0, the formulation above reduces to a simple density-dependent model, and when ε = 1, the formulation above reduces to a density-independent model. The total force of infection on city *j* at time *t* is defined by:

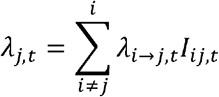

where 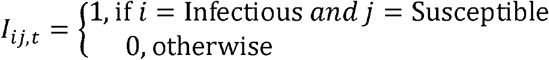

The probability that a susceptible city *j* is invaded at time *t_j_* is

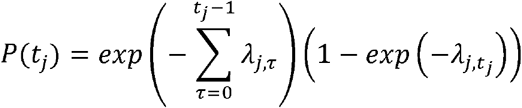

The first part of the equation is the probability that a city escaped invasion from *t* = 0 until just before *t_j_*. The second part is the probability that the city was invaded at *t_j_* given that it was susceptible until that week. The conditional log likelihood is summed over all susceptible cities:

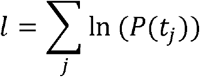

We first investigated null models that only included β. Parameters were then added to test for a spatial effect in transmission, the role of population size of invaded and susceptible cities, and infectivity. Except for β, which is always estimated by Markov chain Monte Carlo (MCMC), parameters can be fixed at 0, at 1 (not γ), or estimated by MCMC. Alternative formulations were also considered with travel time between cities instead of geographic distance (supporting S1 Text).

### Model estimation and computing

Metropolis-Hastings MCMC sampling was used to investigate the posterior distributions of parameters [28, 29]. Because the parameters cannot take negative values, we sampled from a log normal distribution and corrected the Metropolis accept-reject rule for asymmetric jumping. Parameters were updated one at a time. Uniform prior distributions were used for all parameters. Three chains were run for each model with different starting values, and chains were visually checked for convergence after 100,000 iterations with a burn in of 0.2 times the length of the chains (iterations times number of parameters). Median parameter estimates and 95% credible intervals (95% CrI) were calculated from the posterior distributions after excluding the burn in.

DIC was used to compare models. Lower values of DIC are preferred, and a difference of about 5 is considered meaningful [30]. DIC was calculated using the medians of the posterior distributions of the parameters due to non-normality of the likelihood. All analyses were performed in R version 3.5.1.

### Validation of model fit

The probability distribution of the invasion week was calculated for each city based on the observed start of invasion in other cities up to that time. For each virus, this calculation was performed using the median parameter estimates from the posterior distribution. The probability distributions were then compared with the observed invasion weeks.

We also simulated the CHIKV and ZIKV epidemics in Colombia using 1,000 parameter sets sampled from the posterior distribution. For each set, the city (or cities) invaded in the first time step served as the origin of the epidemic. A random deviate was chosen from a uniform distribution between 0 and 1 for each city in each week. If the probability of *t_j_* was higher than the random deviate, the city became invaded. Once invaded, the observed weekly case counts weighted by the generation time distribution were used to model that city’s infectiousness over time.

Epidemic simulations were also used to test the sensitivity of the case count thresholds used to determine the number of invaded cities for each virus.

## Data Availability

Aggregated data and code will be made available on GitHub upon publication in a peer-reviewed journal (https://github.com/kcharniga/zika_chik_invasion).

## Acknowledgements

The authors would like to thank all of the medical and public health professionals involved in the reporting of chikungunya fever and ZIKV disease cases to Sivigila, Colombia’s national public health surveillance system.

## Supporting information captions

**S1 Fig. City elevation**. Comparison of elevation (in meters) between cities that were invaded versus cities that escaped invasion for (a) CHIKV, (b) ZIKV, and (c) CHIKV, ZIKV, or DENV.

**S2 Fig. Example of algorithm used to estimate week of invasion using the generation time method**. (a) The time series for Caucasia in the department of Antioquia during the 2015-2017 ZIKV epidemic. In this figure, week 1 corresponds to the week ending on August 15, 2015, and week 51 corresponds to the week ending on July 30, 2016. The algorithm identifies the point of maximum incidence in the time series and counts backward one week at a time until there are no reported cases. If there are no cases in this week or the prior two or three weeks depending on the infection’s generation time, then this is the week of invasion. If not, the algorithm continues to go back in time until the condition is met. The part of the line in red is the period used to determine the onset of invasion, and the blue dashed line is the estimated invasion week. (b) The same time series as in (a) is shown until the point of maximum incidence (week ending on January 23, 2016). The estimated time of invasion is week 15 rather than week 21 because cases were reported in weeks 16-20.

**S3 Fig. Comparison of estimated invasion weeks using two methods**. A method based on the first reported cases in each city (x-axis) and a method based on the generation time distribution of each infection (generation time method, y-axis) were compared for (a) CHIKV and (b) ZIKV. The black line is y = x. The two methods show good agreement (CHIKV: r = 0.60, ZIKV: r = 0.68).

**S4 Fig. Comparison of estimated invasion weeks using two methods**. A method based on the generation time distribution (generation time method, x-axis) and a piecewise spline method (Charu method, as in [15], y-axis) were compared for (a) CHIKV and (b) ZIKV. 95% confidence intervals are shown for the Charu method only. For some cities, only the point estimate for *t_j_* fell within the 95% confidence interval; this is shown by a lack of vertical bar. The two methods show very good agreement (CHIKV: r = 0.90, ZIKV: r = 0.70).

**S5 Fig. Probability distribution of estimated invasion week (generation time method)**. Colored lines represent the probability distribution of invasion week for (a) CHIKV and (b) ZIKV. The calculations were performed using the median parameter estimates from the posterior distributions of the models using estimated invasion week rather than first reported cases. The black lines show the estimated invasion week in each city using a method based on each infection’s generation time. Values of 0.01 represent probabilities of 0.01 or less.

**S6 Fig. Epidemic invasion simulations (generation time method)**. Simulated invasion for (a) CHIKV and (b) ZIKV from the models using estimated invasion week by generation time method rather than week of first reported cases. Simulated epidemics are shown in light gray. The dark gray lines are the average across the 1,000 simulations. The red lines are the observed incidence curves.

**S7 Fig. Epidemic invasion simulations (all cities)**. Simulated invasion for (a) CHIKV and (b) ZIKV from the models using week of first reported cases and all 1,122 cities in Colombia. Simulated epidemics are shown in light gray. The dark gray lines are the average across the 1,000 simulations. The red lines are the observed incidence curves.

**S8 Fig. Long-distance transmission events**. The distribution of d-D for (a) CHIKV and (b) ZIKV in this study. The dashed blue lines are plotted at the 97.5^th^ percentile (corresponding to 212.00 km and 295.55 km for CHIKV and ZIKV, respectively) and the dashed red lines are plotted at the 99^th^ percentile (corresponding to 344.40 km and 324.52 km for CHIKV and ZIKV, respectively). Long-distance transmission events were defined as invasions that occurred in cities included in the 99^th^ percentile of this distribution.

**S9 Fig. Probability distribution of first reported cases by department for CHIKV**. Black circles are cities that fall within the 95% interval of their expected distribution, and red circles fall outside this distribution. The gray circle in the department of Córdoba represents the city that was invaded in week 0.

**S10 Fig. Probability distribution of first reported cases by department for ZIKV**. Black circles are cities that fall within the 95% interval of their expected distribution, and red circles fall outside this distribution. Gray circles in the departments of San Andrés, Valle del Cauca, and Norte de Santander represent cities that were invaded in week 0.

**S11 Fig. Parameter estimates for CHIKV model fitted using estimated invasion week by generation time method**. The dashed red line shows the median of the posterior distribution of each parameter after removing the burn-in period. The blue line shows the median of the posterior distribution from the model fitted using the method based on first reported cases as in the main text. Only parameters that are estimated in both models are shown.

**S12 Fig. Parameter estimates for ZIKV model fitted using estimated invasion week by generation time method**. The dashed red line shows the median of the posterior distribution of each parameter after removing the burn-in period. The blue line shows the median of the posterior distribution from the model fitted using the method based on first reported cases as in the main text. Only parameters that are estimated in both models are shown.

**S1 Table. Summary statistics of the d-D distributions**. This table shows that ZIKV and CHIKV exhibited similar patterns of transmission. The first six columns have units in km. The seventh column is the total sample size, and the last two columns contain the number of long-distance transmission events for two distance thresholds.

**S2 Table. Estimates of the mean and standard deviation of the generation time distribution**. Estimates were used to calculate city infectivity. All values have units in days.

**S3 Table. Parameter estimates for six models of CHIKV for 337* cities using geographic distance**. Posterior medians and 95% credible interval presented for each parameter. Bold indicates the best-fitting model.

**S4 Table. Parameter estimates for six models of CHIKV for 337* cities using travel time between cities**. Posterior medians and 95% credible interval presented for each parameter. Bold indicates the best-fitting model.

**S5 Table. Parameter estimates for six models of ZIKV for 287* cities using geographic distance**. Posterior medians and 95% credible interval presented for each parameter. Bold indicates the best-fitting model.

**S6 Table. Parameter estimates for six models of ZIKV for 287* cities using travel time between cities**. Posterior medians and 95% credible interval presented for each parameter. Bold indicates the best-fitting model.

**S1 Movie. Monthly chikungunya fever incidence per 100,000 population by department on a hexagonal grid**.

**S2 Movie. Monthly ZIKV disease incidence per 100,000 population by department on a hexagonal grid**. For both movies, the following abbreviations were used for department names: AMA = Amazonas, ANQ = Antioquia, ARA = Arauca, ANT = Atlántico, BOL = Bolívar, BOY = Boyacá, CAL = Caldas, CAQ = Caquetá, CAS = Casanare, CAU = Cauca, CES = Cesar, CHO = Chocó, COR = Córdoba, CUN = Cundinamarca, GUA = Guainía, GUV = Guaviare, HUI = Huila, LAG = La Guajira, MAG = Magdalena, MET = Meta, NAR = Nariño, NDS = Norte de Santander, PUT = Putumayo, QUI = Quindío, RIS = Risaralda, SAA = San Andrés, SAN = Santander, SUC = Sucre, TOL = Tolima, VDC = Valle del Cauca, VAU = Vaupés, VIC = Vichada. Note that the island department of San Andrés is attached to the mainland on the left side of the map.

